# The total number and mass of SARS-CoV-2 virions

**DOI:** 10.1101/2020.11.16.20232009

**Authors:** Ron Sender, Yinon M. Bar-On, Shmuel Gleizer, Biana Bernsthein, Avi Flamholz, Rob Phillips, Ron Milo

**Author notes:** Ron Milo is the corresponding author name here. Equal contribution (Ron Sender, Yinon M. Bar-On).

## Abstract

Quantitatively describing the time course of the SARS-CoV-2 infection within an infected individual is important for understanding the current global pandemic and possible ways to combat it. Here we integrate the best current knowledge about the typical viral load of SARS-CoV-2 in bodily fluids and host tissues to estimate the total number and mass of SARS-CoV-2 virions in an infected person. We estimate that each infected person carries 10^9^-10^11^ virions during peak infection, with a total mass in the range of 1-100 μg, which curiously implies that all SARS-CoV-2 virions currently circulating within human hosts have a collective mass of only 0.1-10 kg. We combine our estimates with the available literature on host immune response and viral mutation rates to demonstrate how antibodies markedly outnumber the spike proteins and the genetic diversity of virions in an infected host covers all possible single nucleotide substitutions.

**Significance:** Knowing the absolute numbers of virions in an infection promotes better understanding of the disease dynamics and the response of the immune system. Here we use the best current knowledge on the concentrations of virions in infected individuals to estimate the total number and mass of SARS-CoV-2 virions in an infected person. Although each infected person carries an estimated 1-100 billion virions during peak infection, their total mass is no more than 0.1 mg. This curiously implies that all SARS-CoV-2 virions currently in all human hosts have a mass of between 100 gram and 10 kilogram. Combining the known mutation rate and our estimate of the number of infectious virions we quantify the formation rate of genetic variants.

---

Estimating key biological quantities such as the total number and mass of cells in our body or the biomass of organisms in the biosphere in absolute units improves our intuition and understanding of the living world (1–4). Such a quantitative perspective could help the current intensive effort to study and model the spread of the COVID-19 pandemic. We have recently compiled quantitative data at the virus level as well as at the community level to help communicate state-of-the-art knowledge about the SARS-CoV-2 virus to the public and researchers alike and provide them with a quantitative toolkit to think about the pandemic (5). Here we leverage such quantitative information to estimate the total number and mass of SARS-CoV-2 virions present in an infected individual during the peak of the infection.

Viral loads are commonly measured in two distinct ways: counting viral RNA genomes by quantitative real-time PCR (qRT-PCR) and measuring the number of infectious units in tissue culture (6). The second approach incubates susceptible mammalian cells with dilutions of a patient sample to determine the amount of sample required to kill 50% of the cells. This value is used to back-calculate the infectious titer in the sample in units of “50% tissue culture infective dose” or TCID_50_ (for example by the Reed and Muench method (7)). The TCID_50_ is analogous (and often quantitatively similar) to the plaque forming units (PFU) assay. Here, we refer to TCID_50_ and PFU more generally as “infectious units”. As these two measurement modalities (RNA genome copies and infectious units) differ in reported values and interpretation -one method measuring the number of RNAs, the other measuring the number of infectious units -we report and compare estimates stemming from both approaches.

To estimate the total number of virions present in an infected individual at the peak of infection, we rely on three studies which measured the concentration of SARS-CoV-2 genomic RNA in the tissues of infected rhesus macaques 2-4 days after inoculation with the virus (8–10). Viral concentrations were measured in samples of all the relevant tissues of the respiratory, digestive and immune systems and values are given in units of genome copies per gram tissue. We use values measured in rhesus macaques as they are the closest organism to humans where such comprehensive data is available. Using these measurements we estimate the total number of virions by multiplying the concentration of viral genomes in each tissue by the total tissue mass (11, 12). We assume that each genome is associated with a virion (i.e, the ratio of virions to genome copies *F*_*virions to RNA copies*_≈1). In case a large fraction of the viral RNA copies are present as “naked” RNA (not encapsulated inside viral particles), using viral RNA copies as a proxy for the number of viral particles could lead to an overestimate. We expand on this source of uncertainty in the discussion. As seen in Figure 1, the lungs are the largest of these tissues on a mass basis (*M*_*lungs*_≈ 1kg). Lungs were also found to harbor the highest concentration of viral RNA (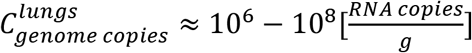, see SI for full details and comparison with additional sources) and we therefore estimate that virions in the lungs are the dominant contributor to the total number of virions in the body during peak infection, with

**Figure 1:**
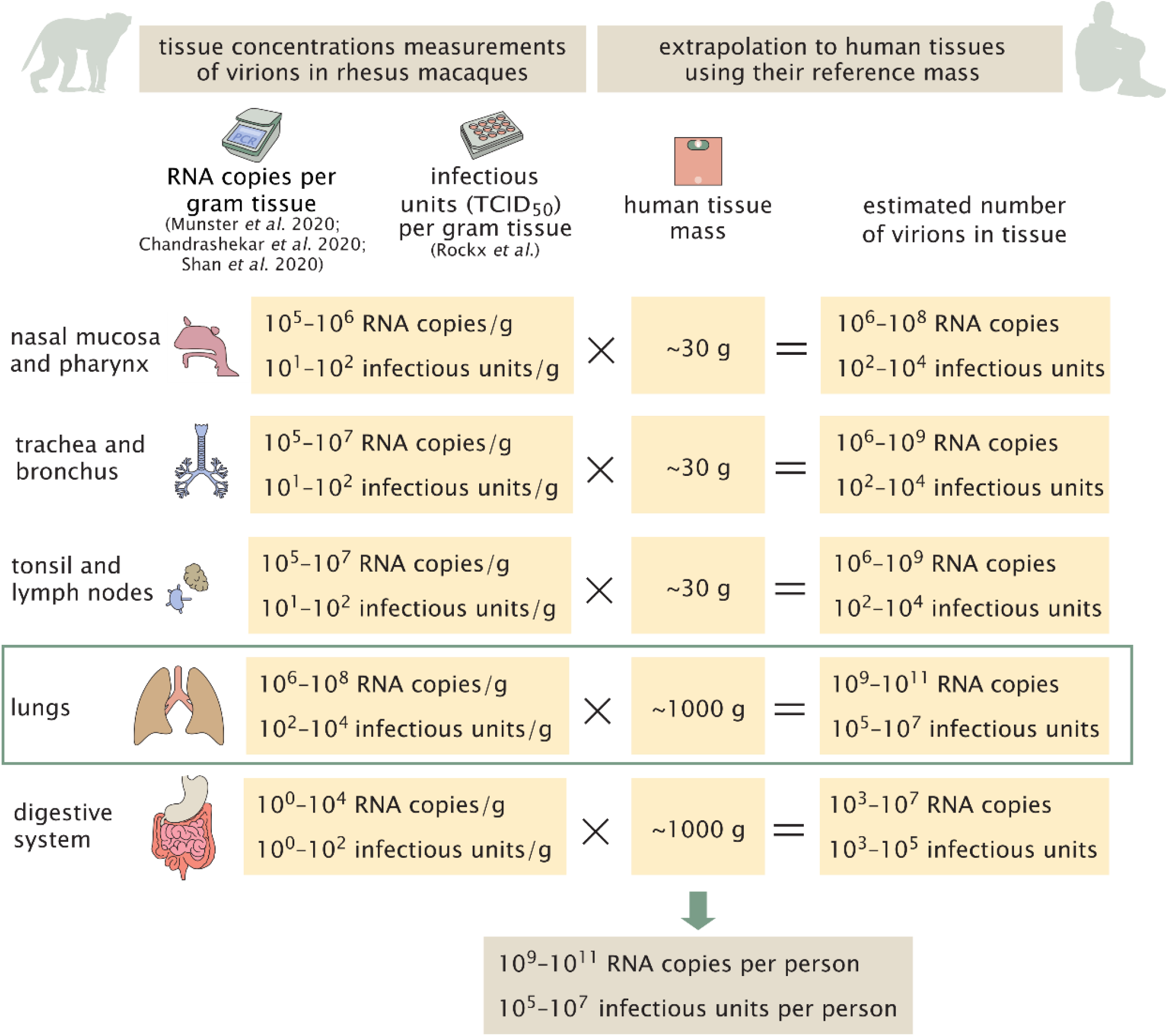
A schematic representation of the estimate of the number of virions in an infected individual. The estimate is made using the viral load measured in a gram of rhesus macaque tissue multiplied by the mass of human tissues in a reference adult person with a total body weight of 70 kg (11). In the digestive tract the concentrations are close to the detection limit.

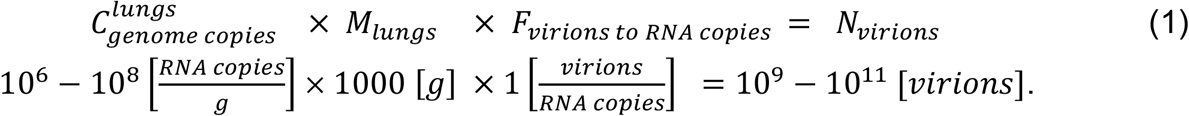

Other tissues, like the nasal mucosa, larynx, bronchial tree and adjacent lymph nodes all have a combined mass of ∼100g (12) and maximal concentrations of 10^6^-10^7^ RNA copies/ml and hence contribute at most an additional 10% to an estimate based solely on the lungs (Figure 1).

Another study (13) measured concentrations of infectious virus in tissues of infected rhesus macaques 4 days after inoculation using cell culture methods. This study reports measurements in units of TCID_50_. The maximal values in these units are much smaller, on the order of 10^3^-10^4^ TCID_50_/ml for lung tissue. Combining these measurements with the volume of adult human lung tissue (≈1L), we get an estimate of 10^5^-10^7^ infectious units in an adult, compared with 10^9^-10^11^ RNA copies, estimated from the other studies (Figure 1). These data suggest a difference of roughly 4 orders of magnitude between RT-PCR measurements of viral RNA and tissue culture measurements of viral titers in TCID_50_ units. To check the consistency of this result with the published literature, we collected 13 studies that measured both SARS-CoV-2 viral RNA copies as well as TCID_50_ or PFU in monkeys and human samples (SI). The characteristic ratio between RNA copy measurements and TCID_50_ measurements is about 4 orders of magnitude but can vary between 3-5 orders of magnitude. We attend to this seeming discrepancy between viral genomic copies and infectious units in the Discussion. We continue to analyze what can be inferred from the evidence that the total number of virions in an infected individual during peak infection is 10^9^-10^11^, and the number of infectious units is 10^5^-10^7^.

While the estimates were performed using a reference value for the lung mass taken from adult men, they can be generalized to the case of women and children. We rely on the multiplication of the viral concentration in the lungs and the total mass of the lungs. Reference values for the lung mass show a value smaller by 20% for women, and 25-75% smaller for children aged 5-15 years (12). Although COVID-19 is known to affect adult men more than women and children (14, 15), there is scarce information regarding difference in viral concentrations across gender and age. One preprint (16) suggests that viral concentration in children is lower by up to an order of magnitude, but the change they measured is not consistent across the entire age range. Assuming the change in measured viral load represents a similar change in viral concentration in the lung tissue, and combining the concentrations with the reduced lung mass, we get that the number of virions in an infected woman is similar to that estimated for men (i.e. of the same order), and that an infected child is probably carrying an order of magnitude less virions.

In addition to analyzing the state of an infected individual during peak infection, we can also estimate the total number of virions and infectious units produced over the course of an infection, as well as the rate of virions production inside a human host. To estimate the total number of virions produced during an infection, we look at its relation to the number of virions at peak infection. The viral load of an infected individual usually increases sharply until reaching the peak and then drops rapidly. The fact that the viral load curve is steep means that a large fraction of all virions produced will be produced close to the peak of infection. The half-life of SARS-CoV-2 virions in the extracellular space was inferred to be about 1-8 hours (17–19). This means that in the 1-3 days near the peak of infection the cumulative viral production needs to be about 3-30 times the observed instantaneous (“snap-shot”) viral load. Because the overall production of virions is dominated by the time period near peak infection, we thus estimate that the ratio between the total production of virions to their peak number is in the range 3-30 (see SI for details, as well as additional derivation of this ratio from observations of infected cells and the residence time of virions in them). Combining this factor with our estimates for the peak number of virions and infectious units we get a total production of 3 ×10^9^-3 ×10^12^ virions, or 3 ×10^5^-3 ×10^8^ infectious units over the complete course of a characteristic infection.

## Calculating the total number of cells infected with SARS-CoV-2

We use our estimate of the total number of infectious units in the body of an infected individual to estimate the number of cells that are infected by the virus during peak infection. In order to estimate the total number of infected cells, we estimate how many infectious units are found in each infected cell as shown in Figure 2.

**Figure 2:**
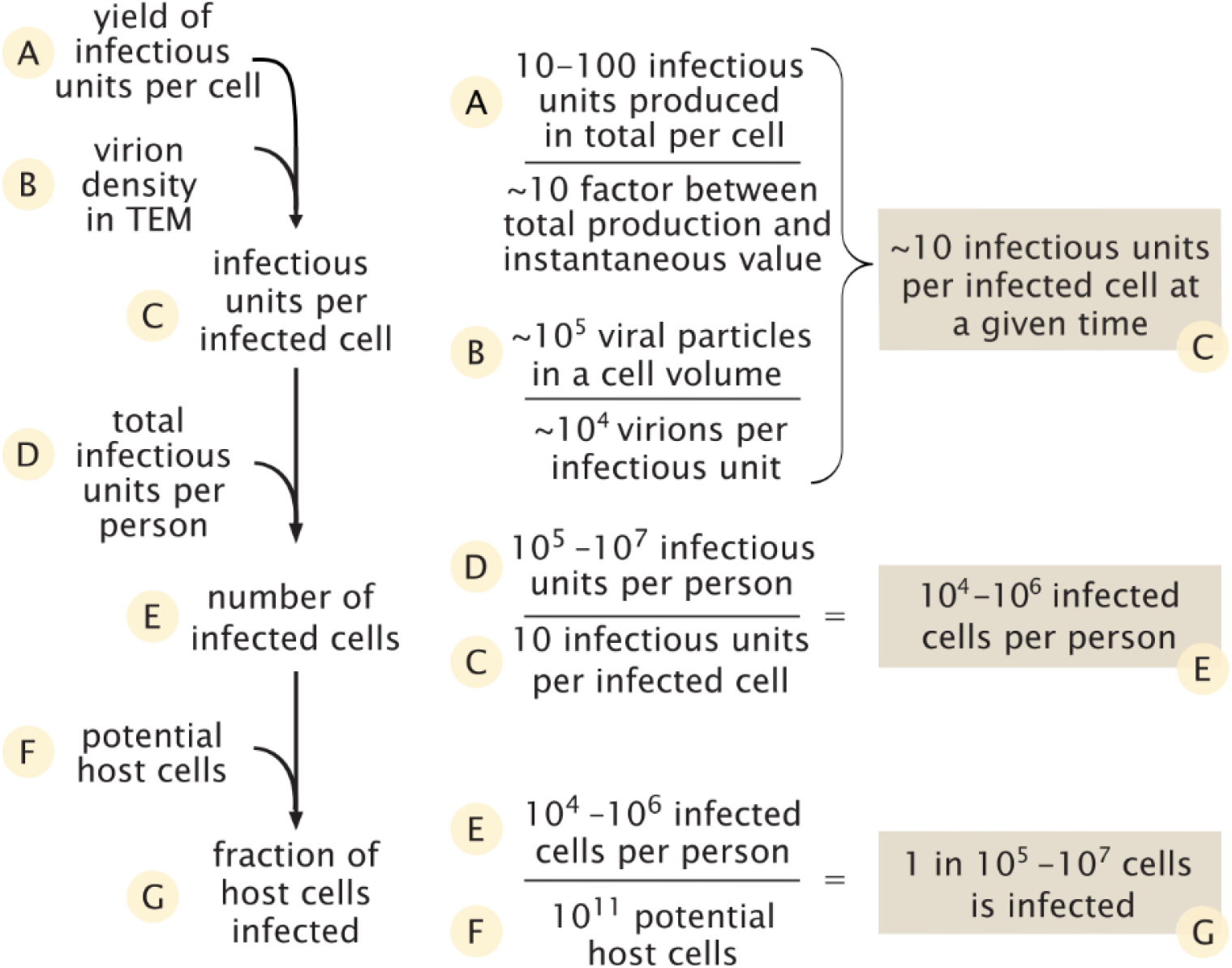
Estimate of the number of infected cells and their fraction out of the potential relevant host cells.

We rely on two lines of evidence in order to estimate the number of infectious units within an infected cell at a given time. The first is data regarding the total number of infectious units produced by an infected cell throughout its lifetime also known as the yield. As we are not aware of studies directly reporting values of the yield of cells infected with SARS-CoV-2, we used values reported for other betacoronaviruses in combination with values we derived from a study (20) of replication kinetics of SARS-CoV-2. Using a plaque formation assay to count the number of infectious units, two previous studies measured the viral yield as either 10-100 or 600-700 infectious units (21, 22). Using reported values for replication kinetics of SARS-CoV-2 (20) we estimated a yield of ∼10 infectious units per cell at 36-48 hours from infection, in agreement with the lower end of these estimates. To convert the total number of infectious units produced overall by a cell into the number of units residing in the cell at a given moment, we estimate the ratio between these two quantities to be 3-30 using two independent methods detailed in the SI. Combining this ratio with our estimate for the total number of units produced by a cell, we thus estimate that, at any given moment, there are somewhere between a few to a few hundreds of infectious units residing in each infected cell.

The second line of evidence concerns the density of virions within a single cell. Several studies have used transmission electron microscopy (TEM) to characterize the intracellular replication of SARS-CoV-2 virions within cells (23–26). Using seven TEM scans taken from those studies we estimated that the density of virions within infected cells is 10^5^ virions per 1 pL (see Dataset S1). As the human cells targeted by SARS-CoV-2 have a volume of ≈1 pL (resulting in a cellular mass of ≈1 ng) (27, 28), TEM data indicate there are ≈10^5^ viral particles within a single infected cell at any point in time. As done above, we assume a ratio of 1 infectious unit resulting per 10^4^ virions. Thus, TEM scans imply that there are ≈10 infectious units that will result from the virions residing inside a cell at any given moment after the initial stages of infection.

Following those lines of evidence we conclude that at a given moment there are ∼10^5^ virions residing inside an infected cell which translates into ∼10 infectious units. Using the ratio of total production to the value at a given time inside the cell, we further conclude that the overall yield from an infected cell is ∼10^5^-10^6^ virions or ∼10-100 infectious units, coinciding with the middle range of measurements from other betacoronaviruses. This estimate also agrees well with recent results from dynamical models of SARS-CoV-2 host infection (29, 30).

We can perform a sanity check using mass considerations to see that our estimate of the number of virions is not beyond the maximal feasible amount. Each virion has a mass of ≈1 fg (5). Hence, 10^5^ virions have a mass of ≈0.1 ng, about 10% of the total mass of a 1 ng host cell and about a third of its dry weight. While a relatively high fraction, this is still within the range observed for other viral infections (31, 32).

Combining the estimates for the overall number of infectious units in a person near peak infection and the number of infectious units in a single cell (*C*_*infectious units per cell*_), we can calculate the number of infected cells around peak infection:

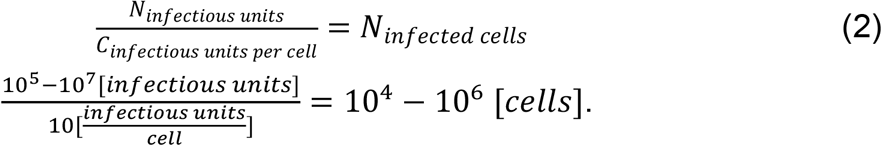

How does this estimate compare to the number of potential host cells for the virus? The best-characterized route of infection for SARS-CoV-2 is through cells of the respiratory system, specifically the pneumocytes (∼10^11^ cells), alveolar macrophages (∼10^10^ cells) and the mucus cells in the nasal cavity (∼10^9^ cells) (27, 28). Other cell types, like enterocytes (gut epithelial cells) can also be infected (33) but they represent a similar number of cells (34) and therefore don’t change the order of magnitude of the potential host cells. As such, our best estimate for the size of the pool of cell types that SARS-CoV-2 likely infects is thus ∼10^11^ cells, and the number of cells infected during peak infection therefore represents a small fraction of this potential pool (1 in 10^5^-10^7^).

## Discussion

Our quantitative analysis establishes estimates for the absolute number of virions present in an infected individual, as well as the number of virions produced during the infection and the total number of infected cells in the body. There are various ways in which one can leverage such quantitative estimates to produce insights regarding COVID-19. First, having absolute estimates allows us to compare them to other quantities in the human body and thus put the number of virions in context and even arrive at new insights. For example, a human body comprises ≈3×10^13^ cells (3). This means that even for our highest estimate, i.e. 10^11^ virions per host, human cells outnumber the virions by more than 100 fold. We can also compare our estimate for the total number of infected cells with the total pool of cells expressing ACE2 (angiotensin-converting enzyme 2) and TMPRSS2 (transmembrane protease, serine 2), the receptor and main protease SARS-CoV-2 relies on for infecting cells. Single-cell RNA-sequencing studies (35–37) indicate that a few percent of the cells in the lungs and airways express ACE2 and TMPRSS2. Most of the cells that have been found to express both are type 2 pneumocytes. While these results might be biased due to drop-out effects in measurements of only a few molecules (37, 38), it is still reasonable that 1%-10% of the lung and airway cells contain the necessary receptor to be infected by SARS-CoV-2, totaling ∼10^9^ cells. This number is several orders of magnitude higher than our estimate for the total number of infected cells during peak infection (10^4^-10^6^). This suggests that out of the cells expressing both ACE2 and TMPRSS2, only a small fraction, e.g. 10^−5^-10^−3^, are infected by the virus.

Because the immune system is the main line of defense against SARS-CoV-2, it is interesting to quantitatively examine the known immune response in comparison with the viral loads we estimated here. For example, we can compare the peak number of viral particles (10^9^-10^11^) to the number of antibodies the body produces to combat SARS-CoV-2 infection. Levels of SARS-CoV-2 specific IgG antibodies (*C*_*IgG*_) were measured 3 weeks after the onset of symptoms, showing a serum concentration of ∼10 µg/mL (39). Only ≈5% of the total anti-spike (the viral protein responsible for allowing the attachment and fusion with the host cell) IgG antibodies has the capacity to neutralize the virus (*f*_*neutralizing*_) (40). Combining the concentration of neutralizing IgG antibodies with a mean IgG molecular weight (*MW*_*IgG*_) of 150 kDa (41) we estimate the number of neutralizing antibodies per mL of serum (*C*_*neutralizing*_):

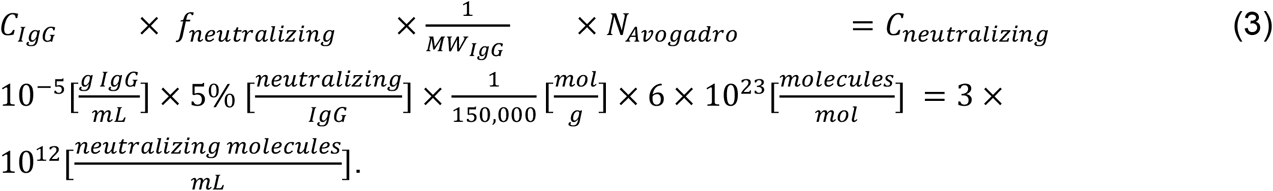

Combining this estimate with the measurement of viral concentration within the lung tissue and accounting for 30-40 spike trimers on each SARS-CoV-2 virion (42, 43) we can estimate the ratio of neutralizing antibodies to viral spike proteins as

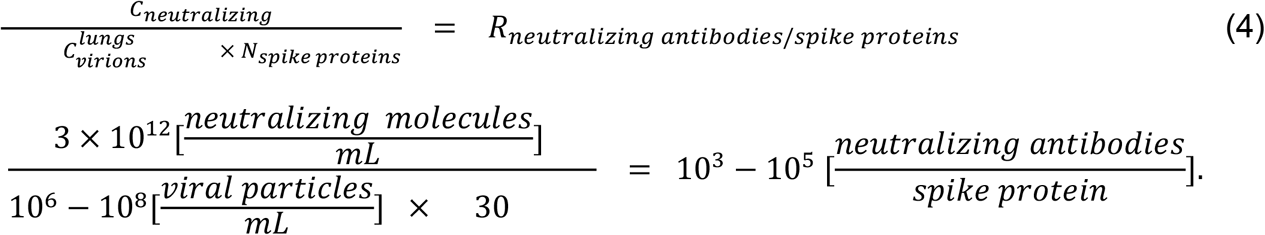

Previous work on other morphologically similar RNA viruses like influenza and flavivirus found that a ratio of 1 bound neutralizing antibody per 2-4 receptor-binding proteins was sufficient to neutralize binding of a virion to its cellular receptor in vitro (44, 45). Taken at face value, our estimate seems to suggest an excess of neutralizing antibodies. There are several factors that will cause the effective concentration of antibodies the virus experiences to be lower. First, the antibody concentrations in the lung tissue tend to be lower than that of the blood. Second, many of the spike proteins are extensively glycosylated. These glycosylations shield many of the binding sites for neutralizing antibodies (43) and thus decrease the efficiency of neutralization (46). However, it is important to remember that the most relevant measure for the effectiveness of antibody neutralization, is the fraction of viral spike proteins that are bound by neutralizing antibodies. This fraction is determined by the strength of the binding of the neutralizing antibodies (nAb) to the viral particles, given by the dissociation constant K_d_ (45). Following the first order relation:

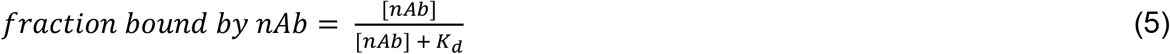

As the dissociation constants for antibody-epitope binding are mostly in the range of 1-10 nM (47, 48) we get:

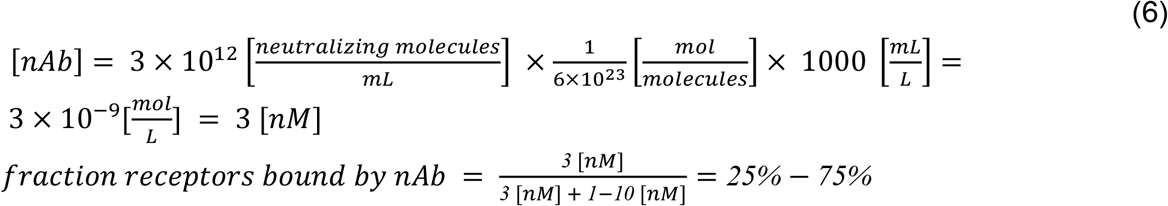

Thus, even though the ratio between the number of neutralizing antibodies and viral particles is high, such a high number of antibodies is essential to ensure that enough of the epitopes are bound (even higher ratio is needed for some antiviral drugs, as shown in the SI).

Beyond the humoral arm of the immune response, T cells are also an integral part of the targeting of viral antigens. Although severe cases of COVID-19 tend to have lower concentration of T cells in the blood, they have a higher fraction of SARS-CoV-2– specific T cells than mild COVID-19 cases (49). Here SARS-CoV-2–specific T cells denotes T cells that showed markers for activation and proliferation after stimulation with SARS-CoV-2 peptide pools (49). We can use the concentrations of CD4+ and CD8+ cells in the blood in combination with their fraction of SARS-CoV-2–specific cells (49) to estimate 1-2 CD4+ cells/μL and 0.2-0.3 CD8+ cells/μL specific for SARS-CoV-2 in convalescent patients and severe cases. Assuming a patient’s blood volume is ∼5L and that 1-2% of lymphocytes reside in the blood (34), we estimate that there are up to 10^9^ SARS-CoV-2 specific T cells in severe cases, with an unknown fraction found in the infected tissue, or 1 per 1-100 viral particles at the peak of infection, and 10^2^-10^4^ such T cells per infected cell.

In our comparisons, we usually rely on our estimates for the characteristic values for the peak viral load in infected individuals, which correspond to the center of the distribution of the measured values (specifically the interquartile range -between the quantiles 25%-75%). However, it is important to note that there is a high degree of variability in viral loads, exceeding 6 orders of magnitude, as can be seen from samples taken from the upper respiratory system (50). This wide variation reflects the difference between people as well as differences in viral load through the progression of infection within an infected individual (51). Thus, extreme cases could exceed the interquartile range provided by an additional two orders of magnitude, reaching values of 10^13^ viral particles in a single person at the peak of infection, while up to 10% of the cells expressing both ACE2 and TMPRSS2 are infected. The variation in the number of virions, as related to the severity of the disease and its outcome, is detailed in the SI. It is also important to note that viral load in different tissues in the host body changes throughout the infection, with some tissues likely infected early on and others later in the infection (52).

Another way in which we can use our estimates to produce new insights is by taking a global view and extrapolating from the numbers observed in a single infected individual to the entire population. For example, we can estimate the number of viral particles residing in all infected humans at a given time. The total number of viral particles at peak infection was shown above to be 10^9^-10^11^ viral particles (this range corresponds to the 25%-75% percentile range). Because the viral loads of individuals are roughly log-normally distributed (53), the arithmetic average of the number of viral particles at peak infection would be on the high end of the range, even beyond the 75th percentile (10^11^-10^12^ particles). There is a rapid drop in viral loads after peak infection, thus the total number of viral particles is dominated by those infected individuals who are close to the infection peak (within 1-2 days). Assuming during most of the course of the pandemic there has been a total of 1-10 million infected people close to peak infection globally at any given time (including those undetected, see SI for details; (54)), we arrive at a total of 10^17^-10^19^ viral particles or 10^13^-10^15^ infectious units at any given time. Similarly, the arithmetic mean of the number of particles produced over the course of infection of an average individual is 10^12^-3 × 10^13^ viral particles 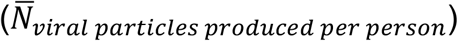, or 10^8^-3 × 10^9^ infectious units (see SI for the detailed derivation of the uncertainty range).

One can contextualize these estimates using an absolute mass perspective. Each virion has a mass of ≈1 fg (5). Therefore even when the body carries 10^9^-10^11^ viral particles, these have a mass of only about 1-100 μg, i.e. 1-100 times less than the mass of a poppy seed. The total mass of virions residing in humanity at a given time is on the order of 0.1-10 kg. Furthermore, using the total number of viral particles produced throughout an infection we can derive the total mass of all the SARS-CoV-2 viral particles ever produced throughout this current pandemic (concentrating on humans which we find to currently dominate over animal reservoirs). We assume the total number of infected people will be in the range of 0.5-5 billion people, representing optimistic and pessimistic future scenarios for the pandemic (see SI for details). To calculate the total number of virions that will have been produced by the end of the pandemic, we multiply the total number of infected people by the total number of viral particles produced over an infection of an average person (which is the arithmetic mean of the distribution across people). We then multiply this number by the average mass of a single virion to find the total mass of viral particles produced globally for such widespread infection (see SI for details of the uncertainty estimate):

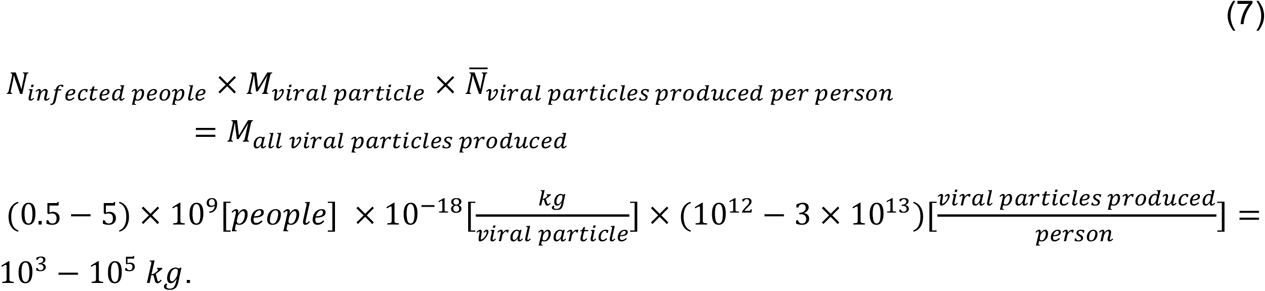

Finally, we use our estimates of the total number of viruses in an infected human to examine the evolution of SARS-CoV-2 and, specifically, estimate the rate of emergence of new variants. When studying the genetic diversity of SARS-CoV-2, we can define two different measures for diversity. The first is the diversity along a genetic lineage of virions -propagating from the ancestral strain in Wuhan until currently circulating virions. The second is the diversity among a population of virions -for example the population of virions present in the body of an infected individual. We start by calculating the average number of mutations accumulated along a specific lineage of ancestor virions leading from the beginning of viral replication in the host until the end of host infection. In these calculations we rely on estimates of the mutation rate per replication cycle per site (3 × 10^−6^ nt^-1^ cycle^-1^) which have been measured for MHV, another betacoronavirus (5). We further assume that each human host is infected by a few infectious units (55–57), and use the estimated yield of ∼10-100 infectious units per cell. Each cycle of infection is therefore assumed to produce 10-100 infectious units that, in turn, go on to infect other cells. As estimated above, there are 3 10^5^-3 10^8^ infectious units produced over the course of an infection. Assuming exponential growth, the entire course of infection will therefore take 3-7 viral replication cycles (Figure 3A). As the SARS-CoV-2 genome has a length of 30,000 nucleotides, we can compute the expected number of mutations accumulating in a virus that is the product of 3-7 replication cycles using the per cycle mutation rate:

**Figure 3:**
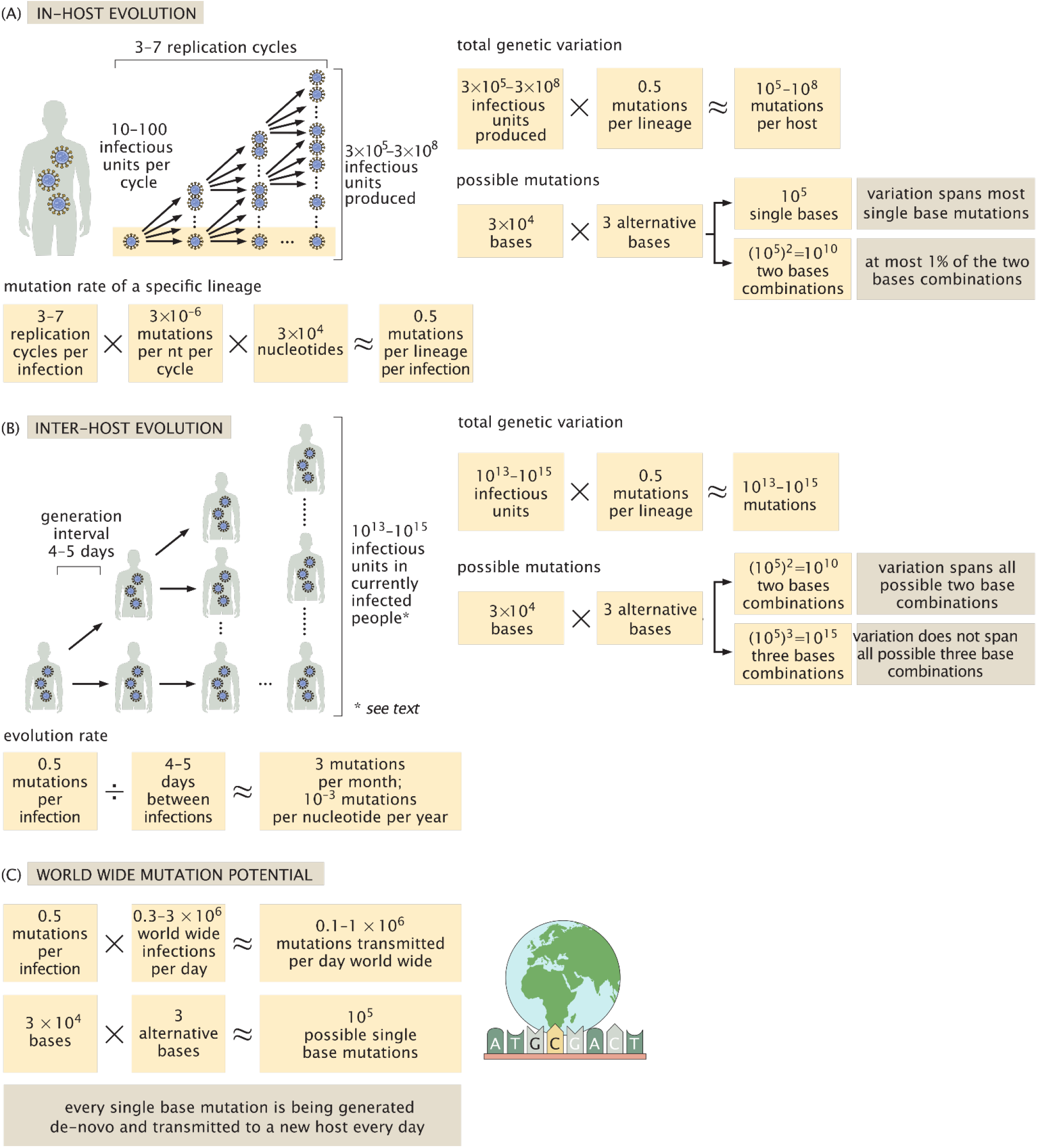
The relationship between the number of virions produced in an infected individual and the evolution of SARS-CoV-2. We use our estimates for the total number of virions produced during an infection, along with other epidemiological and biochemical characteristics of SARS-CoV-2 to estimate the rate of mutation accumulation within an infected host (A) and within the population (B). We consider both the evolution along a specific genetic lineage of virions as well as the diversity among a population of virions -either within an infected host (A) or within the total population (B). In addition, we look at the de-novo mutation generated and transmitted to newly infected in comparison to all possible single base mutations (C).

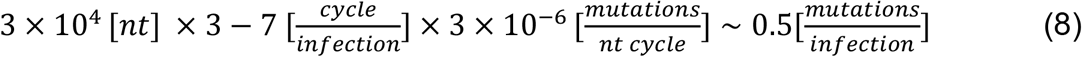

Therefore, if we track a single lineage of virions from the time they started replicating in the body until the end of the infection, this lineage would accumulate in the range of 0.1-1 mutations on average across its entire genome (Figure 3A). Considering that the mean time between successive infections, known as the generation interval, is about 4-5 days, we can estimate an overall rate of ≈3 mutations per month over the course of the epidemic (Figure 3B). This is consistent with empirical values observed during the pandemic for SARS-CoV-2 of about 10^−3^ nt^-1^ yr^-1^ (58, 59), also known as the evolution rate. The evolution rate is estimated from the observed rate of mutation accumulation across sequenced genomes from different time points over the course of the pandemic using reconstruction of phylogenetic trees (59). It therefore includes both the rate of accumulation of neutral mutations and the effects of natural selection. This estimated rate of evolution matches the number of mutations observed in variants present today, about a year after the onset of the pandemic, most of which contain about 20-30 mutations. The extreme examples in terms of number of mutations, of variants such as B.1.1.7, accumulated closer to 40 mutations compared to the first strains isolated.

We can use our estimates of the viral mutation rate to assess the expected rate of appearance of a specific single base mutation. Consider the example of a single nucleotide substitution resulting in the E484K mutation in which the Glutamate (E) in position 484 is replaced with Lysine (K). This mutation requires a specific substitution in a specific location: the first base of the codon must change from G to A. As each nucleotide can mutate to 3 others (e.g. G can become A, T or C) and the genome contains 30,000 nucleotides, there are ≈100,000 possible single nucleotide substitutions to the SARS-CoV-2 genome. As concluded above, about 0.5 mutations are accumulated in every host infection cycle. Without accounting for the effects of selection (i.e. assuming the mutant virions are equally capable of infection and propagation), or the varying chances of mutation among nucleotides, we expect that such a specific mutation will be observed in one out of every ∼200,000 infections. Over the last months, hundreds of thousands of cases have been identified across the world every day and many additional cases have likely gone unidentified. Indeed, as shown in Figure 3C, the estimated number of mutations generated daily (10^5^-10^6^ mutations/day) likely exceeds the total number of possible single nucleotide substitutions to the SARS-CoV-2 genome (≈10^5^ substitutions) assuming 0.3-3 million new cases a day worldwide. As such, our estimates imply that every single base mutation is being generated de-novo and transmitted to a new SARS-CoV-2 host, somewhere in the world, every day.

In addition to considering a specific lineage of SARS-CoV-2 viruses, we can also consider the genetic diversity at the population level and estimate the total variability across the entire repertoire of infectious units produced during a single course of infection. As we estimated that 3 × 10^5^-3 10^8^ infectious units are produced during an infection, each one resulting from a lineage of ancestors and mutations, we expect overall to have about 10^5^-10^8^ mutations across all of the infectious units. Some of these mutations that occured in early cycles will appear in many later progeny within the host, while those generated in the most recent cycle will appear in only one viral genome. Because the SARS-CoV-2 genome is 30,000 nucleotides long, the 10^5^-10^8^ mutations across all of the virions produced over the course of a single infection probably cover every possible single nucleotide substitution (Figure 3A). They even cover a significant fraction of the possible pairs of single nucleotide substitutions. If we look globally at the entire number of infectious units of SARS-CoV-2 currently present within the infected human population, which we estimated above at 10^13^-10^15^, we expect that every combination of two nucleotide substitutions and many, though not all, three nucleotide substitutions will be present in at least one infectious unit (Figure 3B).

This large genetic diversity might naively imply that advantageous mutations will rapidly take over the population due to natural selection, but there are several factors which slow down the rate of selection. These factors include epistasis, a phenomenon where a single mutation becomes advantageous only on the background of other specific mutations. Another key factor is the genetic bottleneck imposed during the transmission of virions between infected individuals. These bottlenecks are expected to slow selection as only a tiny fraction of the diversity generated in the host is passed onto future generations (55–57). This quantitative understanding brings to focus cases in which selection can occur for a significant amount of time with no bottlenecks, such as the case of long and persistent infections, for example in immunocompromised patients (60–62). We thus conclude that careful accounting of the number of virions can give insight into the process of viral evolution within and across hosts.

One of the strengths of a holistic quantitative analysis such as the one performed here is its ability to expose interesting “quirks” that are otherwise elusive. One such observation is the ratio of ∼10^4^ between the RNA copies measured using RT-PCR and the number of infectious units measured in TCID_50_. Ratios on the order of 10^3^-10^4^ between viral particles and PFUs were observed in animal viruses such as Poliovirus and Papillomavirus (63). Naively, such a ratio would suggest that only 0.01% of the virions produced are actually infectious. This ratio implies that SARS-CoV-2 is not very efficient in producing infectious progeny. While we do not have a clear explanation for this seeming low efficiency, there are several possible factors that will affect this ratio. First, measuring RNA copies may not correspond directly to actual virions, but also measures naked viral RNA. Second, while TCID_50_ is the most widely available assay for measuring infectious titer, it may not accurately reflect the actual number of infectious virions, for example, because conditions in the assay may not be optimal for SARS-CoV-2 infection. Another possibility is that many virions are non-infectious due to the neutralizing effect of binding antibodies, and thus the ratio may represent the effect of the immune response, and change over the period of infection.

Beyond exposing these quantitative aspects, a holistic analysis allows us to identify major knowledge gaps in the available literature. For example, the virion yield per infected cell is known only from a few studies on different kinds of betacoronavirus from over 40 years ago (21, 22). Similarly, measurements of the mutation rate per nucleotide per cycle in SARS-CoV-2 are of much interest but missing. As discussed above, the quantitative relationship between viral RNA copies, viral particles and infectious units is not fully characterized for SARS-CoV-2, and thus further research could help better constrain and explain the differing values. In addition, a model describing the quantitative relationship between antibody production and infection metrics would help quantitatively test the estimates presented here.

Establishing estimates for the total number and mass of SARS-CoV-2 virions in infected individuals allows us to connect together various aspects of the pandemic, from immunology to evolution, and to highlight emerging patterns and relationships not obviously evident. Having better quantitative information on the process of infection at the cellular level, the intra-host level and the inter-host level will hopefully empower researchers with better tools to combat the spread of COVID-19 and to understand its evolution, including the rise of variants of concern.

## Data Availability

To generate our estimates of cellular turnover, we extracted values from the literature as detailed in the spreadsheet files available as supplementary information

## Acknowledgments

We would like to thank Itai Benhar, Gidon Eshel, Shai Fuchs, Thierry Mora, Eran Segal, Maya Shamir, Ziv Shulman, Huicheng Shi, Harinder Singh, Einat Vitner, Aleksandra Walczak, John Yin for valuable feedback on this manuscript.

## Funding

This research was supported by the European Research Council (Project NOVCARBFIX 646827), Israel Science Foundation (Grant 740/16), Beck-Canadian Center for Alternative Energy Research, Dana and Yossie Hollander, Ullmann Family Foundation, Helmsley Charitable Foundation, Larson Charitable Foundation, Wolfson Family Charitable Trust, Charles Rothschild, Selmo Nussenbaum, Miel de Botton (R.M.), the National Institutes of Health (1R35 GM118043-01 (Maximizing Investigators Research Award)) (R.P.), Merkin Institute for Translational Research (R.P.), the Israeli Council for Higher Education (CHE) via the Weizmann Data Science Research Center and by a research grant from Madame Olga Klein – Astrachan (R.S.). R.M. is the Charles and Louise Gartner Professional Chair. Y.M.B. is an Azrieli Fellow

## Notes

### Competing Interest Statement

The authors have declared no competing interest.

### Summary of Updates

Additional analysis of mutation rate and few corrections. Text improvements. Supplementary information addition.

