## Supplementary material for "The total number and mass of SARS-CoV-2 virions": SI Appendix

PNAS
www.pnas.org

**Supplementary Information for**

### The total number and mass of SARS-CoV-2 virions

Ron Sender[1†], Yinon M. Bar-On[1†], Shmuel Gleizer[1], Biana Bernsthein[1,2], Avi Flamholz[3], Rob Phillips[3,4], Ron Milo[1]

[1]Weizmann Institute of Science, Rehovot 7610001, Israel
[2]Present address: Ragon Institute of MGH, MIT and Harvard, Cambridge, MA 02138, USA
[3]California Institute of Technology, Pasadena, CA 91125, USA
[4]Chan Zuckerberg Biohub, 499 Illinois Street, SF CA 94158, USA

[†] Equal contribution
*Ron Milo is the corresponding author name here


**This PDF file includes:**

Supplementary text
Figures S1 to S2
Tables S1 to S2
Legends for Dataset S1
SI References

**Other supplementary materials for this manuscript include the following:**

Dataset S1
Jupyter notebooks JN1-JN2, available at https://gitlab.com/milo-lab-public/number_of_virions

#### Supplementary Information Text

In the following we provide a detailed account of the methodology used for estimating the viral load at peak infection, as well as some additional analyses. All the data used to generate our estimates, along with the code for analyzing the data, are open-source and available in the accompanying Dataset S1 spreadsheet file, (its legend is given below) and at https://gitlab.com/milo-lab-public/number_of_virions. Our analysis pipeline consists of two additional Jupyter notebooks, which use the data extracted from the literature as input for generating our estimates for the viral concentrations in the different tissues, and for derivation of the uncertainty of our analysis results.

##### Viral load at peak infection

We estimate the number of virions at peak infection by multiplying the viral concentration inside the different tissues by their reference mass in an adult male. In order to estimate the viral concentration in different tissues we refer to models of rhesus macaques, which enable direct measurements of viral concentration in the homogenized tissue, at necropsy. A thorough literature search for the terms: “rhesus macaque” + ”SARS-CoV-2”, using Google Scholar produced 15 papers, of which 11 contained measurements of tissue specific viral concentrations.

Longitudinal measurements of viral load (in nasal swabs or bronchoalveolar lavage) in these 11 studies show that peak infection is usually reached a few days after inoculation. Thus, we further focused on measurements from three studies (1–3) taken 2-4 days from inoculation with the virus. A comparison with measurements from additional 6 studies (4–9) taken at 5-7 days after inoculation showed a small decrease in viral load, that may be caused by measurements taken after the peak infection of the animals (Figure S1).

The lung tissue was the dominant contributor to the overall number of viral genome copies, having the largest mass and viral concentrations of the tissues considered. Here we walk through our methodology for estimating viral loads in lung tissue. First, for each animal, we calculated the mean viral load across several locations in the lungs. We then took the geometric mean of mean viral loads from different animals to arrive at our estimate for the viral load at peak infection. We arrive at a final estimate of 6.5±0.5 $\log_{10}$ viral RNA genomes per gram lung tissue as measured by qRT-PCR. However, this average represents only a small sample of 9 animals, and thus it is preferable to also consider the variation across measurements taken from various locations of the lungs of those animals (58 measurements). The interquartile range (IQR) of these measurements, representing the range between the 25 percentile and the 75 percentile, is 5.5-7.6 $\log_{10}$ viral RNA per gram tissue. Comparison of nasal swabs taken from human and rhesus macaques indicate that viral loads tend to be 0.5-1 orders-of-magnitude lower in rhesus macaques. Therefore, we consider the range of 6-8 to be representative of the $\log_{10}$ viral concentration in human lungs, given in units of RNA copies per gram tissue.

We only found two papers reporting tissue culture measurements of infectious units in $TCID_{50}$ (4, 10), one which took measurements 4 days after inoculation (10). We use the

IQR of their measurements (2.3-3.5 $TCID_{50}$/g), and rounded it to the closest order of magnitude to conclude a reference range of 2-4 for the $\log_{10}$ of the infectious units concentration in $TCID_{50}$ units (figure S1 panel C).

**The ratio between the total number of virions produced over an infection and the peak number of virions**

In order to estimate the ratio between the total number of virions produced over an infection and the peak number of virions we apply two methodologies. First, we look at the "micro-level", concerning an infected cell. Second, we look at the "macro" level, using results from dynamic models of in-host infection. On the micro-level, we can look at the ratio between the number of virions residing in a single infected cell at a given moment, and the total number of virions the infected cell will yield over its infection. As the in-host dynamic indicates that most of the cells are infected in parallel, at the peak of infection, the two ratios are approximately equal. This ratio is dependent on the average residence time of the virions inside the cells ($\tau_{residence}$), and on the duration of the infection of the cell ($T_{infected}$). It takes SARS-CoV-2 about a day to infect a cell and produce new virions (11) and the average infection time of the cell is 1-2 days (12, 13). The residence time of virions within the cells is not known, so we assume that it is in the range of a few hours (corresponding to the clearance time of virions, in the extracellular space, see details below) to a maximum of a half a day, close to the entire duration of infection of the cell. The ratio between the yield ($Y_{total\ production}$) and the number of virions residing in the cell at a given moment ($C_{virions}$) is given by:

$$\frac{Y_{total\ production}}{C_{virions}} = F_{micro} = \frac{T_{infected}}{\tau_{residence}}$$

$$F_{micro} = \frac{1-2\ days}{0.1-0.5\ days} = 2-20$$

On the macro level, we look at studies of in-host viral kinetics. For simplicity we focus on a simple model of exponential growth and decay (14). Following this model, we can get an analytical form for the number of virions ($V$) as a function of the time since infection ($t$):

$$V(t) = \begin{cases} V_0 e^{\alpha t} & 0 \le t \le t_p \\ V_{max} e^{-\beta t} & t_p \le t \le t_f \end{cases}$$

Where $V_0$ is the number of virions at the time of infection; $V_{max}$ is the maximal number of virions; $t_p$ is the time in which the virion number is maximal; $t_f$ is the time in which the number of virions drops below detection level; $\alpha, \beta$ are the exponential growth and decay and they can be expressed as functions of the other parameters:

$$\alpha = \frac{ln(\frac{V_{max}}{V_0})}{t_p};\ \beta = \frac{ln(\frac{V_{max}}{V_0})}{t_f - t_p}.$$

The overall production of virions is therefore given by the area under the curve of $V$, multiplied by the clearance rate of virions $c$. The area under the curve ($S_V$) can be analytically calculated by integration of $V$ over the time of infection:

$$S_V = \int_0^{t_f} V(t)dt = \int_0^{t_p} V_0 e^{\alpha t} dt + \int_{t_p}^{t_f} V_{max} e^{-\beta t} dt = \frac{V_{max} - V_0}{\alpha} + \frac{V_{max} - V_0}{\beta}$$
$$= \frac{V_{max} - V_0}{ln(\frac{V_{max}}{V_0})} t_f$$

We can now get an expression for the overall ratio between total production and peak number of virions:

$$\frac{Y_{total\ production}}{C_{virions}} = F_{macro} = \frac{S_V c}{V_{max}} = \frac{V_{max} - V_0}{V_{max}\ ln(\frac{V_{max}}{V_0})} c t_f$$

As $V_{max}$ is much larger than $V_0$ we get that in approximation:

$$F_{macro} = \frac{c t_f}{ln(\frac{V_{max}}{V_0})}$$

We estimated $V_{max}$ to be in the range of $10^9$-$10^{11}$. Assuming that each human host is infected by a few infectious units (15–17), $V_0$ is in the range of $10^2$-$10^4$.
Hence $ln\left(\frac{V_{max}}{V_0}\right) \approx 10 - 20$. The average duration of infection $t_f$ is in the range of 10-15 days (14). Viral clearance rate was estimated in several in-host modeling studies to be in the range of 2-100, while most of the values were in the range of 4-30 (18–20) corresponding to a half-life in the range of 0.5 hour to few hours (matching estimates for other viruses such as Zika (21) and influenza (22)). Combining those values we get an estimate for the ratio in the range of 3-30.
We therefore conclude that based on the two methods, the ratio between the overall production of virions and the maximal number of virions is in the range 3-30.

**The possible connection of the variation in the total number of virions to the severity of the disease and its outcome**

The large variation between individuals also raises the question of how the total viral load is correlated to the severity of disease. A recent meta-analysis of viral loads (23) shows that although the duration of viral shedding is shorter for asymptomatic patients on average, the average viral loads of asymptomatic patients are similar to those of symptomatic patients. Thus, we expect there would be no order-of-magnitude difference in the peak number of viral particles for asymptomatic patients, but the overall production of virions would be smaller. Regarding the viral load profile among severe and critical patients there is inconclusive evidence concerning the average viral concentrations. Some sources indicate that their viral load is not significantly different from those of patients with mild cases (23, 24) while others find a substantial difference (25). However, there is strong evidence indicating that the duration of viral shedding is longer for patients with severe cases, so we expect the total production of virions to be larger in those cases. Furthermore, a comparison by clinical outcome shows that non-survivors have on average lower cycle threshold levels of 6 Ct units (26), a difference that can be translated to a factor of ~60-fold higher viral concentration. This indicates that non-survivors have on average 1-2 orders of magnitude higher number of viral particles and infected cells.

**Derivation of the uncertainty distribution of the results using Latin Hypercube Analysis**

As discussed in the text, viral concentrations are best described by a log-normal distribution. The parameters of the log-normal distribution characterizing the distribution of viral loads across individuals have an uncertainty associated with them, which should be propagated to the final estimate of the total number of viruses produced during an infection of an average individual. In addition, three other variables that are part of the derivation of the final result, each one with associated uncertainty which must also be propagated:

1. <u>The ratio between the number of viral particles in a host at peak infection and the total number of virions produced over the entire course of infection</u>. As shown above we estimated this ratio to be in the range of 3-30. Therefore, we approximated the uncertainty of that variable by a log-uniform distribution with the range of 3-30.
2. <u>The number of infected people that are near their peak infection at a given time</u>. We estimate this number by taking the daily number of newly identified cases worldwide, which, for most of the epidemic, was in the range of 100,000-500,000 (27). Multiplying by a factor of 2-4, representing the number of days for which viral loads are near-maximal, and by another factor of 3-5 to account for unidentified cases. Thus, we assume there are between a million and ten million infected individuals near peak infection at a given moment. We approximated this uncertainty as a log-uniform distribution with the range of $10^6$-$10^7$.
3. <u>The predicted cumulative number of infected people at the end of the pandemic</u>. As the span of possible future dynamics of the epidemic is immense we tried to approach it by finding reasonable bounds following two extreme scenarios. The pessimistic scenario is one in which humanity emerges from the pandemic only when it reaches herd immunity, after infecting for example 60-70% of the human population. Assuming the fraction of reinfections is small we can use an upper bound for the overall number of infected people as ~$5 \times 10^9$. The optimistic extreme scenario is one in which humanity emerges from the pandemic within one year thanks to a massive vaccination effort. In this scenario we optimistically assume a relatively low infection rate of 0.1-0.3 million per day as a representative rate for that period, and that including the number of unidentified cases this value is about double (though it may also be much higher). Hence we estimate that by the end of the year 200-300 million new cases would have accumulated on top of a similar number of cases counted so far (with about 100 million of them having been identified). Hence, in this optimistic scenario we reach a total of ~$0.5 \times 10^9$ infected cases before the end of the pandemic. Using

these to bounds we thus approximated the uncertainty in the cumulative number of infected people by a log-uniform distribution with the range of 0.5-5x$10^9$

In order to combine the uncertainty in the estimates of the different variables into conclusive results we use the Latin Hypercube Analysis (LHA) method. This method is similar to Monte-Carlo analysis and is performed by sampling independently multiple times from the distribution of each of the variables. However, in the process of LHA sampling, the distributions are pre-divided into bins and sampled without replacement (such that each sample is taken from another bin), ensuring a better representation of the whole distribution given a limited number of samples. Figure S2 depicts a sampling from four variables that define the distribution of viral particles at peak infection and the total mass of particles that would be produced over the epidemic. Panels A-C present the sampling process that ensure that every bin of the distribution is represented, and panel D present the posterior distributions.

**Comparison of viral concentration and antiviral drug concentration**

The total number of virions and their tissue concentrations can help us understand the mechanism of action of antiviral drugs, such as Remdesivir. In their study with rhesus macaques Williamson et al. (7) measured a median concentration of ≈3 nmol of the activated drug within 1 gram of homogenized lung tissue after six days of treatment. This is ≈$10^{15}$ molecules per 1 gram of tissue, about 7-8 orders of magnitude more than the concentration of virions within the tissue. It appears that this ratio of antiviral drug to viral concentration is needed as the reported concentrations are also close to the measured EC50 (50% effective concentration) of ≈10 $\mu$M measured in tissue culture (28) To convert measured EC50 to units of molecules per gram, we use a tissue density ($\rho_{tissue}$ ) of ≈1 g per mL:

$$EC_{50} \times N_{Avogadro} \times \rho_{tissue} = C_{mulecules}$$

$$10^{-5}[\frac{mol}{L}] \times 6 \times 10^{23}[\frac{molecules}{mol}] \times \frac{1}{1000} [\frac{L}{g}] \approx 10^{16}[\frac{molecules}{g}].$$

**Area encompassed by peak viral particles**

Beyond the mass-based perspective, the size of the SARS-CoV-2 virions can also serve to give an intuition for the number of virions that reside in a patient. The size of a viral particle can be approximated by a sphere with 100nm diameter, such that puting 100 virions in a square side by side would cover an area of 1µm x 1µm. Repeating the same exercise for all $10^9$-$10^{11}$ viral particles residing in an infected person at peak of infection, they would cover an area of about 1cm x 1cm. The total surface area of the lungs' alveoli is about ≈100 $m^2$ (29), hence putting all the SARS-CoV-2 viral particles side by side would cover only about a millionth of it.

#### Supplementary Figures

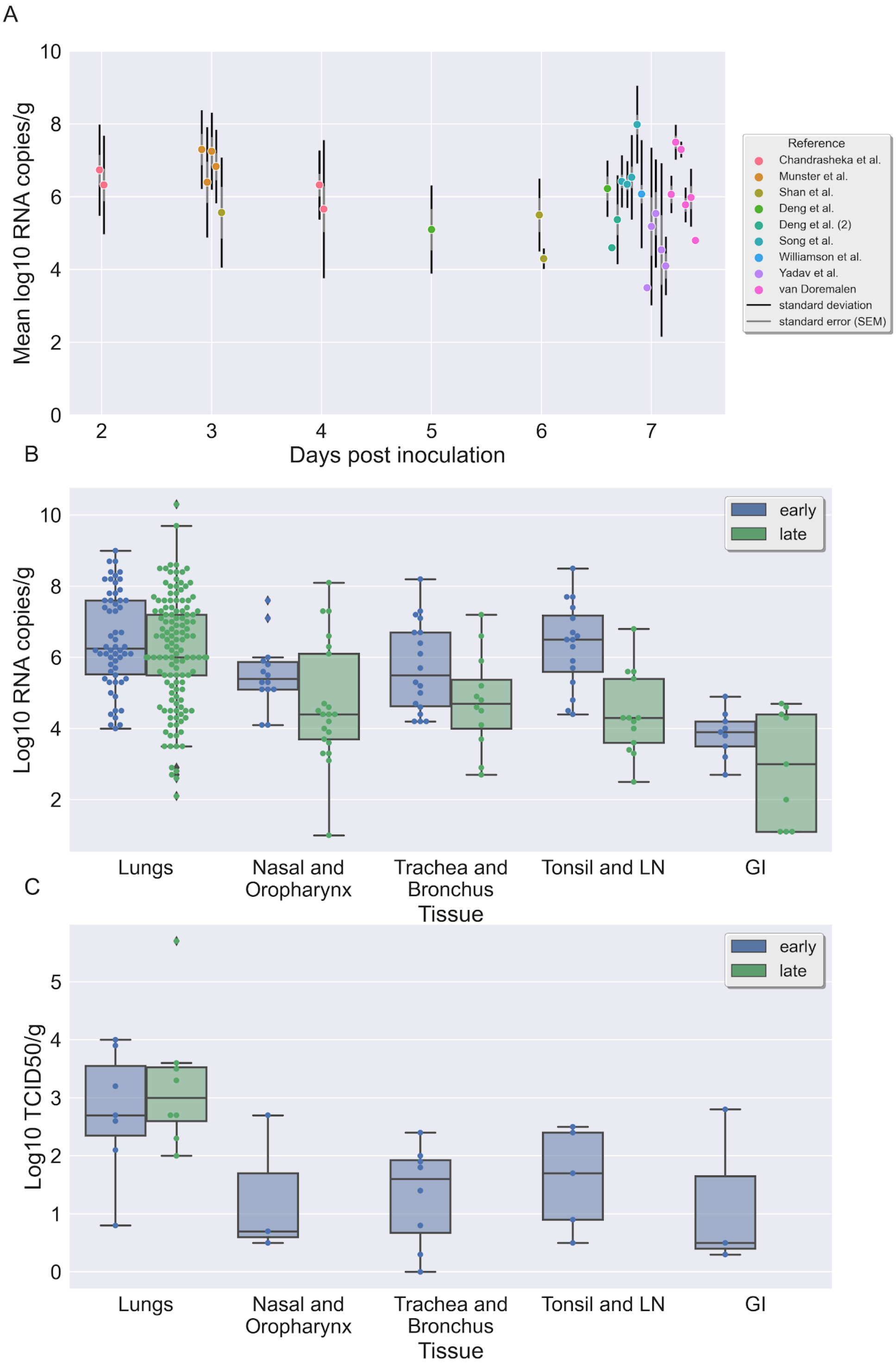


**Fig. S1. The viral concentration in tissues of rhesus macaques**. A. Scatter plot of the log10 of viral concentrations, given in RNA copies per gram versus the day of the measurement at necropsy (measured from inoculation). Each of the points represent the average of all measurements taken from one animal (except from the one representing Williamson et al., for which the data is given for all 6 animals participating). The error bars represent the standard deviation (black) and standard error of the mean (grey) for each animal. B. Boxplot and swarmplot of the distribution of all measurements of the log in base 10 of measurements of RNA copies per gram, given in 9 studies, divided by tissue and whether the measurement was taken more than 4 days after inoculation. C. Boxplot and swarmplot of the distribution of all measurements of the log in base 10 of measurements of $TCID_{50}$ per gram, given in 2 studies, divided by tissue and whether the measurement was taken more than 4 days after inoculation.

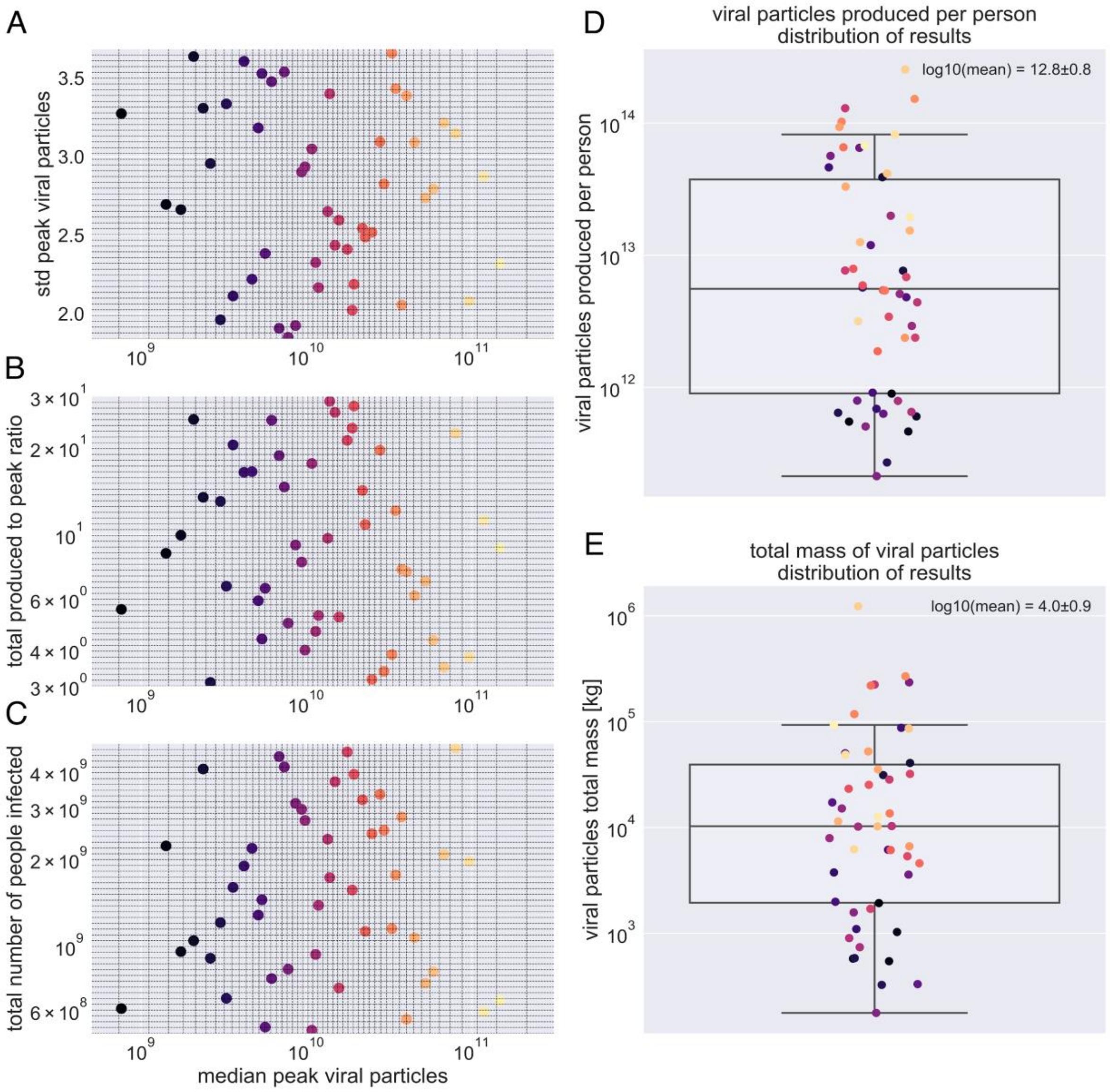


**Fig. S2. Derivation of the uncertainty range for the average number of viral particles produced per person and the total mass of viral particles produced over the epidemic, using Hypercube Latin Analysis**. Panels A-C present 50 points sampled randomly with the Hypercube Latin Analysis method, such that for each of the four variables (median peak viral particles, standard deviation of peak viral particles, the ratio between the total production and the number of particles at the peak, the number of total people infected) there is a representation 2% quantile (1/50) of the distribution. Panel D presents the resulting distribution for the number of virions per person produced over an infection. Panel E presents the distribution of total mass of all the virions that would be produced throughout the epidemic. The labels present the mean of the distribution of log10 of the values. The color of the point corresponds to the order of sampling, such that each color represents the same sample in the five panels.

#### Supplementary Tables

**Table S1. Summary of viral concentration measured in rhesus macaques tissues, from the literature.** The measurements from 11 studies were summarized and divided by tissue and by the time the sample was taken (at necropsy). Statistics for measurements in units of log10 RNA copies/g are given both for the entire dataset of measurements, and for aggregation by specific animals. Those in log10 TCID/g are given only for all measurements, as there was no separation in the main source (10). The reference ranges for the tissue were estimated based on the interquartile range, rounded (up in most cases) to the near order of magnitude.

std = standard deviation; SEM = standard error of the mean; IQR= interquartile range (percentiles 25-75).

[#] Many of the measurements from the gastrointestinal tract were negative, but omitted in this statistics. The lower bound of this range is thus supposed to be the limit of detection.

| **tissue** | **sample time [≤4 dpi]** | **log10 RNA copies/g** | | | | | | **log10 TCID/g** | | |
|---|---|---|---|---|---|---|---|---|---|---|
| | | **by measurements** | | | **by animal** | | | **by measurements** | | |
| | | **mean ± std** | **IQR** | **n** | **mean ± SEM** | **IQR** | **n** | **mean ± std** | **IQR** | **n** |
| **Lungs** | early | 6.5 ± 1.4 | **5.5 - 7.6** | 58 | 6.5 ± 0.5 | 6.1 - 6.8 | 9 | 2.8 ± 1.1 | **2.4 - 3.6** | 7 |
| | late | 6.2 ± 1.5 | **5.5 - 7.2** | 154 | 5.8 ± 0.3 | 4.9 - 6.3 | 27 | 3.2 ± 1.2 | **2.6 - 3.5** | 8 |
| **Nasal mucosa and oropharynx** | early | 5.5 ± 1 | 5.1-5.9 | 21 | 5.5 ± 0.3 | 5 - 6 | 8 | 1.3 ± 1.2 | **0.6 - 1.7** | 8 |
| | late | 4.7 ± 1.7 | 3.7-6.1 | 14 | 4.6 ± 0.4 | 4 - 4.7 | 10 | - | **-** | 0 |
| **Trachea and Bronchus** | early | 5.7 ± 1.2 | **4.6 - 6.7** | 21 | 6.1 ± 0.5 | 5 - 7.1 | 9 | 1.3 ± 0.9 | **0.7 - 1.9** | 8 |
| | late | 4.9 ± 1.3 | **4.2 - 5.6** | 14 | 4.6 ± 0.4 | 3.7 - 5.4 | 14 | - | **-** | 0 |
| **Tonsil and Lymph nodes** | early | 6.3 ± 1.1 | **5.5 - 6.9** | 19 | 6.4 ± 0.4 | 5.7 - 6.8 | 8 | 1.6 ± 0.9 | **0.9 - 2.4** | 5 |
| | late | 4.2 ± 1.1 | **3.5 - 4.3** | 11 | 4.3 ± 0.6 | 3.5 - 4.5 | 6 | - | **-** | 0 |
| **Gastro-intestinal tract** | early | 2.2 ± 2.1 | **0 - 4**[#] | 16 | 2.7 ± 0.7 | 0 - 3.6[#] | 8 | 1.2 ± 1.4 | **0 - 1.7**[#] | 3 |
| | late | 2.9 ± 1.6 | **0 - 4.4**[#] | 9 | 2.9 ± 0.5 | 0 - 3[#] | 3 | - | **-** | 0 |

**Table S2. Literature review for the ratio between viral genome copies and infectious virions measured in $TCID_{50}$.** Summary of data that was collected from 13 studied, according to different organisms and types of samples.
* Each of six studies measuring isolated strain (30–36) had provided only one estimate. The mean and standard deviation was calculated aggregating all 6 data points.

| Organism | Sample type | Reference | mean log10 of ratio RNA copies/ infectious units | standard deviation log10 of ratio RNA copies/ infectious units |
|---|---|---|---|---|
| **African green monkeys** | BAL | (37) | 4.9 | 1.5 |
| | Nasal swab | | 4.6 | 0.7 |
| | Throat swab | | 4.7 | 0.6 |
| | Vero E6 cell line | (36) | 4.2 | 0.2 |
| | **Total** | | **4.5** | **0.9** |
| **Rhesus macaques** | Lung tissue | (4) | 2.4 | 0.6 |
| | BAL | (1, 7) | 4.5 | 1.2 |
| | Nasal swab | (7) | 5.0 | 2.0 |
| | Rectal swabs | | 2.8 | 2.2 |
| | Throat swabs | | 4.1 | 1.6 |
| | **Total** | | **4.0** | **1.8** |
| **Human** | Isolated strain | (30–35) | 4.2* | 0.8* |
| | Calu-3 cell line | (36) | 4.3 | 0.2 |
| | Constructed airway tissue | | 4.1 | 0.3 |
| | Nasal epithelial cells | (38) | 2.9 | 0.8 |
| | Nasal swab | (39) | 4.2 | 0.5 |
| | **Total** | | **3.8** | **0.9** |
| **Total** | | | **4.2** | **1.1** |

**Dataset S1 (separate file).**

The S1 Data spreadsheet includes several tables that give details for the resources collected and calculations made throughout this study. Each tab provides calculations regarding one subject. Sheets are divided to several groups, as follows:

- Total viral particles currently - estimation of the average number of viral particles in an infected host and derivation of the total mass of viral particles in currently infected population.
- Viral particles in an infected person - estimation of the number of viral particles using tissue viral concentration measured in rhesus macaque (in both RNA copies and $TCID_{50}$) and tissue mass (29, 40).
- Gender & age effects - possible effects of age and gender based on the tissue driven estimate and viral load variation in human samples (41).
- Infected cells estimate - estimation of the number of virions within cells and the total number of infected cells.
- Analysis of TEM images - estimation of the number of viral particles using TEM images from several sources. In order to estimate the number of viral particles residing in an infected cell.
- Antibodies - Comparison of antibody levels to viral particles concentrations and calculation of the fraction of bounded neutralizing antibodies.
- Tissue conc. (RM) summary - Statistics of viral concentration in tissues of rhesus macaques both in $TCID_{50}$ and RNA copies. Derived from the database below, using Python jupyter notebook.
- Tissue conc. (RM) dataset - Extracted from the various sources, of samples taken from the tissue of rhesus macaques at necropsy.
- $TCID_{50}$ vs. RNA - review - Literature review of the ratio between SARS-CoV-2 concentration measured in $TCID_{50}$ and RNA copies
- Antiviral drug - antiviral drug concentration measured in rhesus macaque tissues by Williamson et al. (7)
